# Dysgraphic features in motor neuron disease: a review

**DOI:** 10.1101/2021.01.31.21250861

**Authors:** Edoardo Nicolò Aiello, Sarah Feroldi, Alice Naomi Preti, Stefano Zago, Ildebrando Marco Appollonio

## Abstract

**Background:** Motor neuron disease (MND) patients can show oral language deficits mimicking those of frontotemporal degenerations (FTD). Although dysgraphic features have been also reported within the MND-FTD *continuum*, their characteristics and clinical relevance are still largely unexplored.

**Aims:** To profile writing disorders in MND patients can help further define their cognitive semiology and thus conveys relevant clinical entailments. Therefore, this study aimed at reviewing evidence of writing impairment in MND patients. This review was implemented and reported by consulting Preferred Reporting Items for Systematic Reviews and Meta-Analyses guidelines. Qualitative/quantitative measures of writing abilities in MND patients was the primary outcome. Both group studies and case reports/series were taken into consideration. Twenty-four contributions were included out of an initial *N*=83. Potential biases in generalizing results were qualitatively controlled for by extracting background, disease-related, neuropsychological and neuroanatomofunctional secondary outcomes.

**Main Contribution:** Fifteen studies assessed writing abilities in Japaneses patients, whereas the remaining eight in western patients. Central dysgraphic features were reported in both neuropsychologically-impaired and –unimpaired MND patients. Phonetic/phonological paragraphias and morpho-syntactic errors were frequently reported. Although FTD was frequently co-occurent, neither cognitive nor language impairment fully accounted for writing impairment in some patients. By contrast, evidence of peripheral dysgraphia was scarce. Patients displaying writing deficits often presented with bulbar signs and perisylvian cortices involvement (including Exner’s area and the left angular gyrus). Writing deficits proved to be associated with abnormalities in executive functioning and its neural substrates. Writing-to-dictation tasks as well as writing samples assessment proved to be useful to detect writing errors.

**Conclusions:** Dysgraphic features in MND patients might be due to dysfunctions of the graphemic buffer – and possibly the phonological route. The lexico-semantic route appeared to be less involved. However, a mixed peripheral/central involvement cannot be ruled out. In this population, executive/attentive deficits are likely to contribute to writing errors as well. Writing deficits might thus be specific of MND patients’ cognitive/language impairment profile. The evaluation of writing abilities *via* writing-to-dictation/narrative writing tasks may be useful when assessing cognition/language in both neuropsychological-impaired and -unimpaired MND patients - especially when severe dysarthria/anarthria is present and prevents clinicians from assessing oral language.

## 1. Introduction

Motor neuron diseases (MND) are currently regarded as multi-system disorders affecting both motor and *extra*-motor systems [Al-Chalabi *et al*., 2016]. Due to both physiopathological and genetic similarities shared with frontotemporal degenerations (FTD) [Burrell *et al*., 2016], up to 50% of MND patients present with cognitive and/or behavioral deficits within the FTD *spectrum* [Pender *et al*., 2020].

Advances in cognitive semiology of MND patients are relevant to identify sensitive markers of cognitive impairment, which negatively influences their multi-disciplinary clinical management and their ecological functioning [Christidi *et al*., 2018; Huynh *et al*., 2020].

Language deficits mimicking those of primary progressive aphasias (PPA) [Gorno-Tempini *et al*., 2011] are estimated to be moderately prevalent in MND patients (35-40%) [Strong *et al*., 2017]. Language dysfunctions in MND patients affect phonological, lexico-semantic and morpho-syntactic components, as well as connected speech and frontally-mediated communicative functioning [Pinto-Grau *et al*.,2018].

Although language in MND patients has been mainly investigated in the oral modality, writing deficits have been also reported [Pinto-Grau *et al*.,2018] - mostly in Japanese patients, whilst rarely in western languages [Ichikawa *et al*., 2012]. The features of acquired writing disorders in MND patients are nonetheless still largely unknown.

The differences between the Japanese writing system and those of western languages – with respect to both linguistic aspects and neuro-cognitive underpinnings [Sakurai, 2019] – make it challenging to integrate available evidence. Indeed, as opposed to western writing systems that are mostly alphabetic, Japanese written language encompass both a phonogram- (*kana*) and an ideogram-based (*kanji*) system – which underlie different neuro-cognitive mechanisms and can be thus selectively damaged [Ichikawa *et al*., 2012; Sakurai, 2019]. If compared to alphabetic systems, *kana* words can be paralleled by words with a regular phoneme-to-grapheme correspondence (and are thus more likely to be processed *via* a phonological route), whereas *kanji* words by irregular words - as having their phonetics/phonology associated with their semantics (being thus more likely to be processed *via* a lexico-semantic route) [Sakurai *et al*., 1997; Purcell *et al*., 2011; Ichikawa *et al*., 2012; Sakurai, 2019]. As far as *kana* and *kanji* writing is concerned, both generalized and selective deficits have been frequently reported in neurological disorders affecting perisylvian language networks [Sakurai, 2019]. Therefore with regard to central writing disorders, to adapt western nosography (*e*.*g*., phonological *vs*. surface dysgraphia) to Japanese writing deficits is not trivial [Sakurai *et al*., 1997]

To profile writing disorders in MND patients is also diagnostically relevant since: a) the detection of language deficits is sufficient in order to classify those patients as cognitively impaired [Strong *et al*., 2017]; b) writing is useful to assess language functioning in severely dysarthric/anarthric patients [Ichikawa *et al*., 2012].

Aim of the present work was thus to review evidence reporting writing impairment in MND patients in order to draw cross-linguistically valid inferences regarding its features.

## 2. Methods

### 2.1. Search strategy

Preferred Reporting Items for Systematic Reviews and Meta-Analyses guidelines [Moher *et al*.,2009] were consulted in order to implement and report the present review. The following search terms were entered into Scopus and PubMed databases on October 19^th^, 2020: “amyotrophic lateral sclerosis” OR “motor neuron disease” in combination with (AND) “writing error*” OR “writing disorder*” OR “writing dysfunction*” OR “writing deficit*” OR “writing error*” OR “dysgraphi*” or “agraphi*”. Fields of search were the title, the abstract and the key-words in Scopus, whereas the title and the abstract in PubMed.

### 2.2. Inclusion criteria and outcomes

For a contribution to be included, a) writing abilities had to be assessed (*via* either qualitative or quantitative methods) b) in MND patients. The following primary outcomes were extracted: features of writing impairment; writing assessment methods; *kana* and/or *kanji* errors (for Japanese contributions); neural correlates of writing abilities. Review and meta-analysis were excluded. Both group studies and case reports/case series, as well as both longitudinal and cross-sectional studies, were taken into consideration. The following secondary outcomes were extracted (when applicable): patients’ demographics (age, sex, education, language) and clinical features (phenotype [Chiò *et al*., 2011], disease duration, presence of bulbar signs, motor-functional outcome [Cedarbaum *et al*., 1999]); neuropsychological (NPs) *vs*. motor onset [Mioshi *et al*., 2014]; presence of dementia (and type); presence of cognitive and/or behavioral impairment (CI/BI; and type); features of oral language impairment (LI); neural measures.

A qualitative method of writing assessment (*e*.*g*., evaluation of everyday-life writing samples) was assumed if not otherwise stated. A motor onset was assumed if there was not an explicit statement regarding cognitive/behavioral deficits as the presenting symptoms. When a generic diagnosis of dementia was posed, behavioral variant of FTD (bvFTD) was assumed if NPs deficits appeared to fall under those described in Rascovsky *et al*.’s (2011) diagnostic *criteria*. Neural correlates of writing abilities were taken into account only when correlations with writing measures were explicitly investigated.

### 2.3. Bias assessment

The vast majority of the aforementioned secondary outcomes [see 2.2] were taken into account in order to minimize potential biases in generalizing results.

With regard to demographics, since writing abilities are learned and sharpened during school years, low levels of educational attainment might lead to a misinterpretation of writing difficulties. Moreover, taking into consideration different languages might prevent from inferring cross-linguistically generalizable conclusions.

Regarding neurological outcomes, the occurrence of bulbar signs (and thus possibly dysarthria/anarthria) might lead to an overestimation of writing disorders (if present) since language functioning can be solely/are preferably assessed in the written modality [Cobble, 1998].

From a NPs standpoint, both the occurrence of *extra*-linguistic CI/BI and co-morbid non-aphasic dementia might contribute to writing disorders [Graham, 2000]. Therefore, it was of interest to determine whether deficits in domains other than writing abilities could to an extent account for dysgraphic features [Ardila & Surloff, 2006].

### 2.4. Study selection process

Study selection process is shown in Figure 1. *N*=76 contributions were identified through the first level search strategy and were screened within the title and abstract; among them, *N*=37 were excluded based on inclusion criteria. The remaining *N*=39 were selected for further eligibility - along with *N*=7 contributions that were identified through hand-search and judged as being likely to meet inclusion criteria.

After removing duplicates and again applying inclusion criteria, *N*=22 contributions were excluded and *N*=23 were selected for inclusion.

*N*=4 contributions were available in Japanese, and were thus included by only taking into account the abstract and possibly other sections available in English.

**Figure 1.**
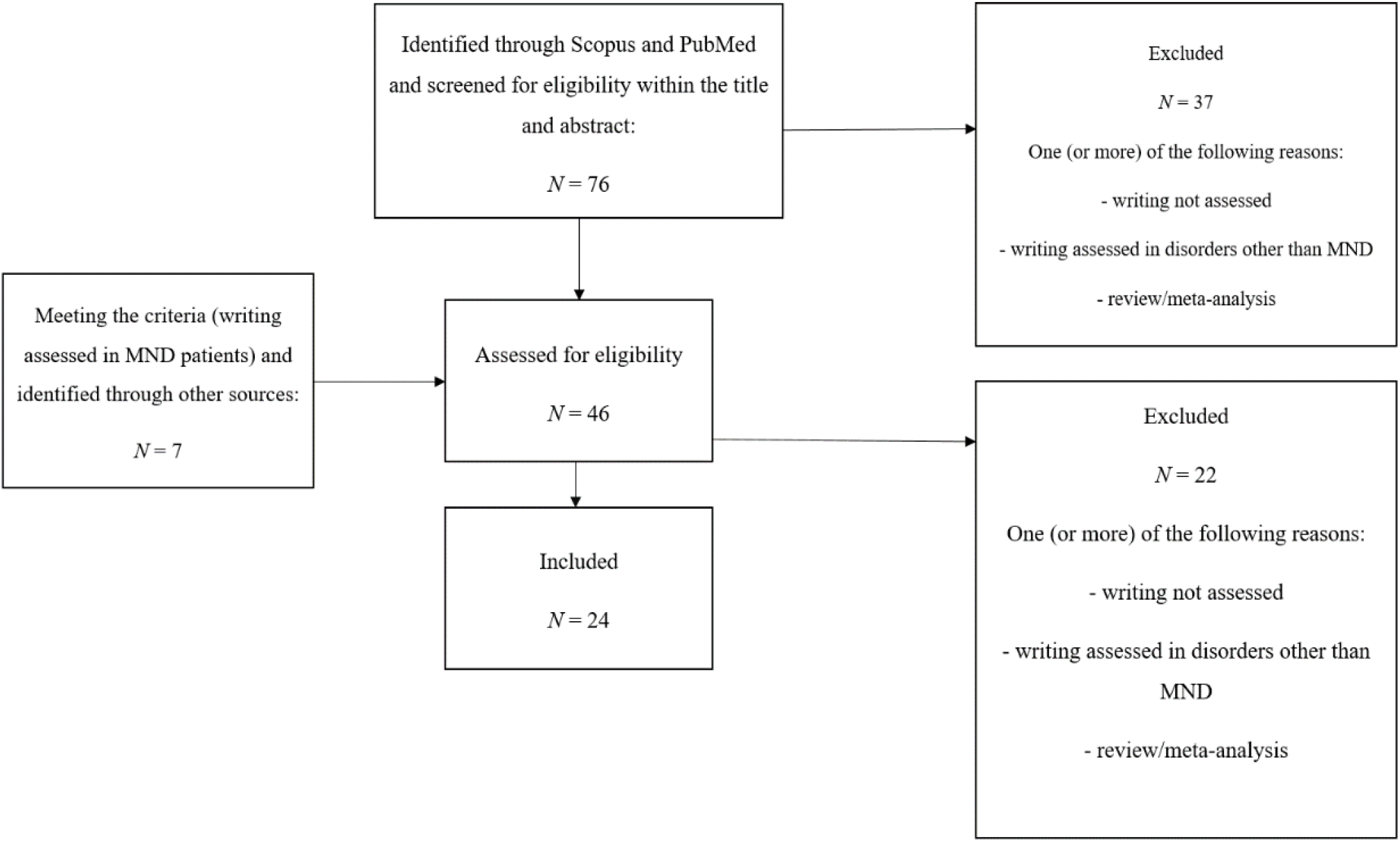
PRISMA-inspired flow-chart showing study selection process. **Notes**. PRISMA=Preferred Reporting Items for Systematic Reviews and Meta-Analyses; MND=motor neuron disease.

## 3. Results

### 3.1. Outcomes overview

Extracted outcomes are summarized in Table 1; an overview of background variables is reported in Table 2. Only *N*=5 contributions were group studies, whereas all the remaining were either case reports or case series. The majority of studies investigated writing abilities in Japanese patients; the remaining assessed western patients. The most prevalent phenotype across studies was amyotrophic lateral sclerosis (ALS); several studies adopted the generic label “MND”. Patients with bulbar signs were highly represented. The vast majority of studies included patients showing either dementia or NPs impairment, whereas only 4 studies reported unimpaired patients.

**Table 1.** Overview of background outcomes.

| *N*<br>**pts.** | Languages | Age<br>(y.; *M* and<br>*SD range*) | Sex<br>(F/M) | Education<br>(y.; *M* and *SD range*) | Disease duration<br>(mo.; *M* and *SD range*) | Phenotype<br>(*N stud.*) | ALSFRS-R<br>(0-48; *M* and *SD range*) | Bulbar signs<br>(*N stud.*) | FTD/<br>NPs deficits<br>(*N stud.*) | NPs/mixe onset<br>(*N stud.*) |
| --- | --- | --- | --- | --- | --- | --- | --- | --- | --- | --- |
| ≈178 | **JA.:16**<br>**IT.:2**<br>**EN.:3**<br>**FR.:1**<br>**SV.:1**<br>**BG.:1** | 41-75<br>7-12.2 | 81/97 | 5-18<br>2.13-4.3 | **ALS/MND:**<br>12.3-168<br>5.1-36<br>**PLS:**<br>49.2-102<br>24-36 | **ALS:**14<br>**MND:**6<br>**PLS:**2<br>**PBP:**1<br>**LMND:**1 | 25.3-41.7<br>4.2-8.4 | 20 | 14/8 | 10 |
| **Notes.** Abbreviations: pts.=patients; y.=years; EN.=English; FR.=French; IT.=Italian; SV.=Swedish; BG.=Bulgarian; mo.=months; F=female; M=male; stud.=studies; MND=motor neuron disease; ALSFRS-R=ALS Functional Rating Scale - Revised. Summarizations comprise available data only. |  |  |  |  |  |  |  |  |  |  |

**Table 2.**
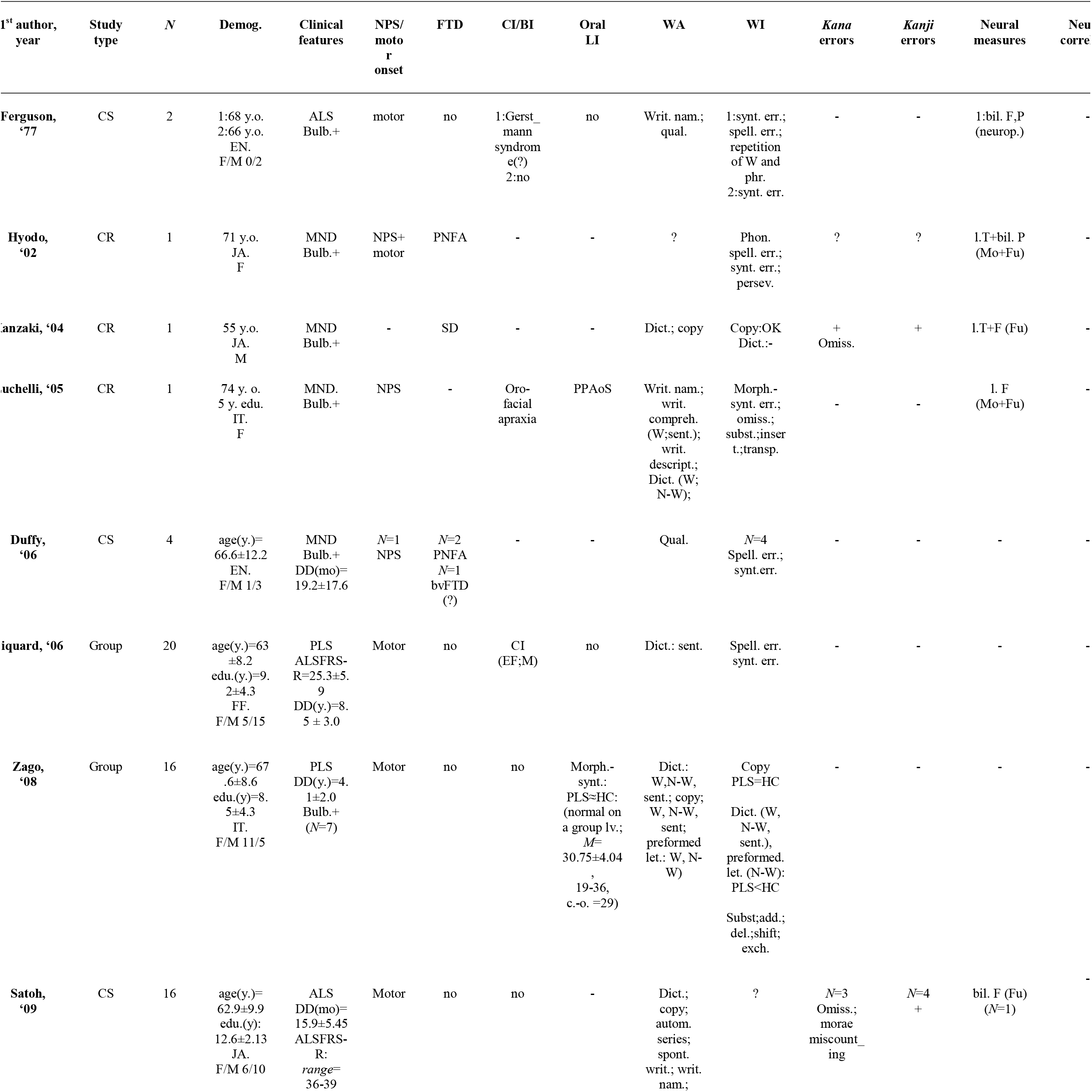
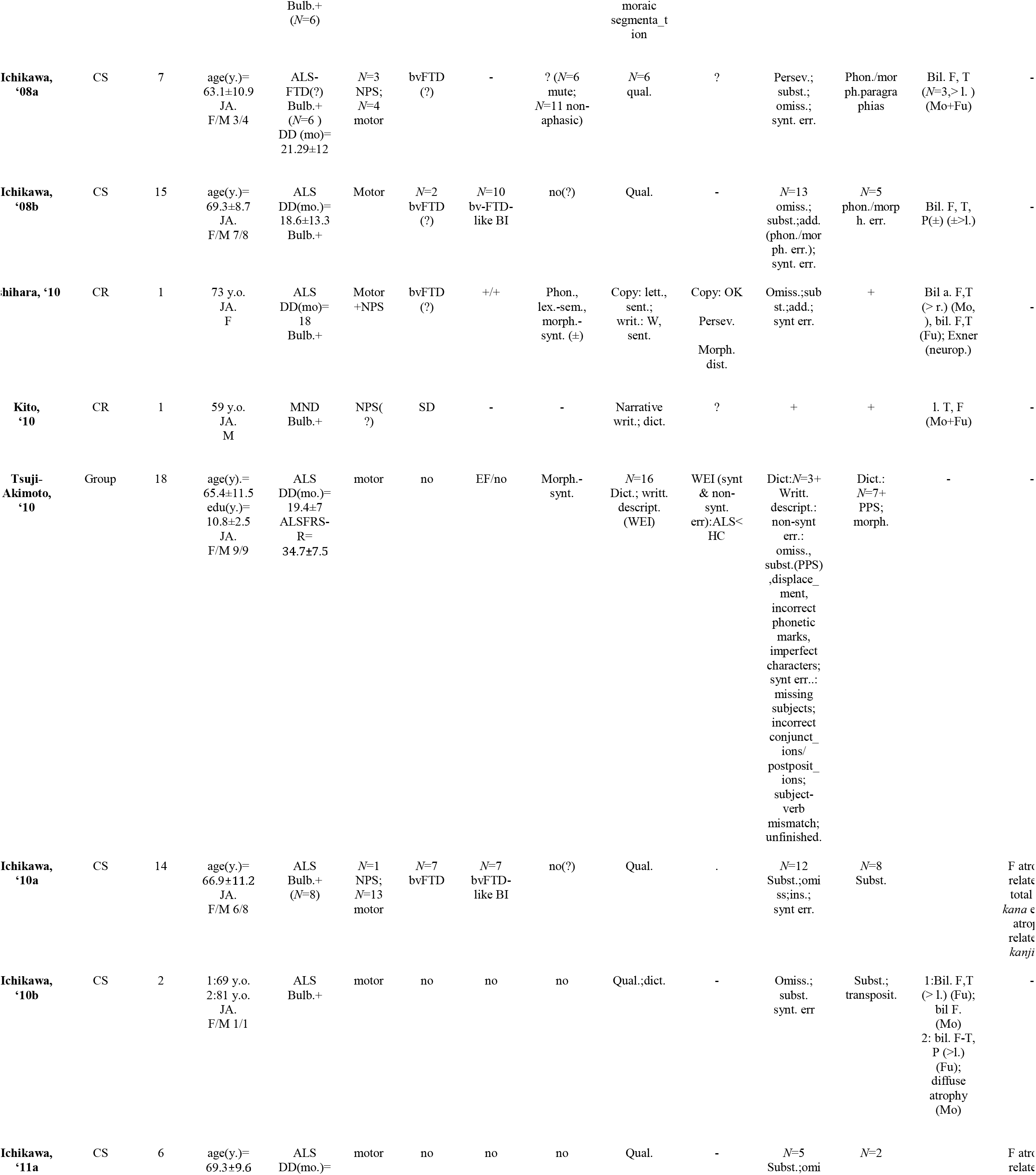
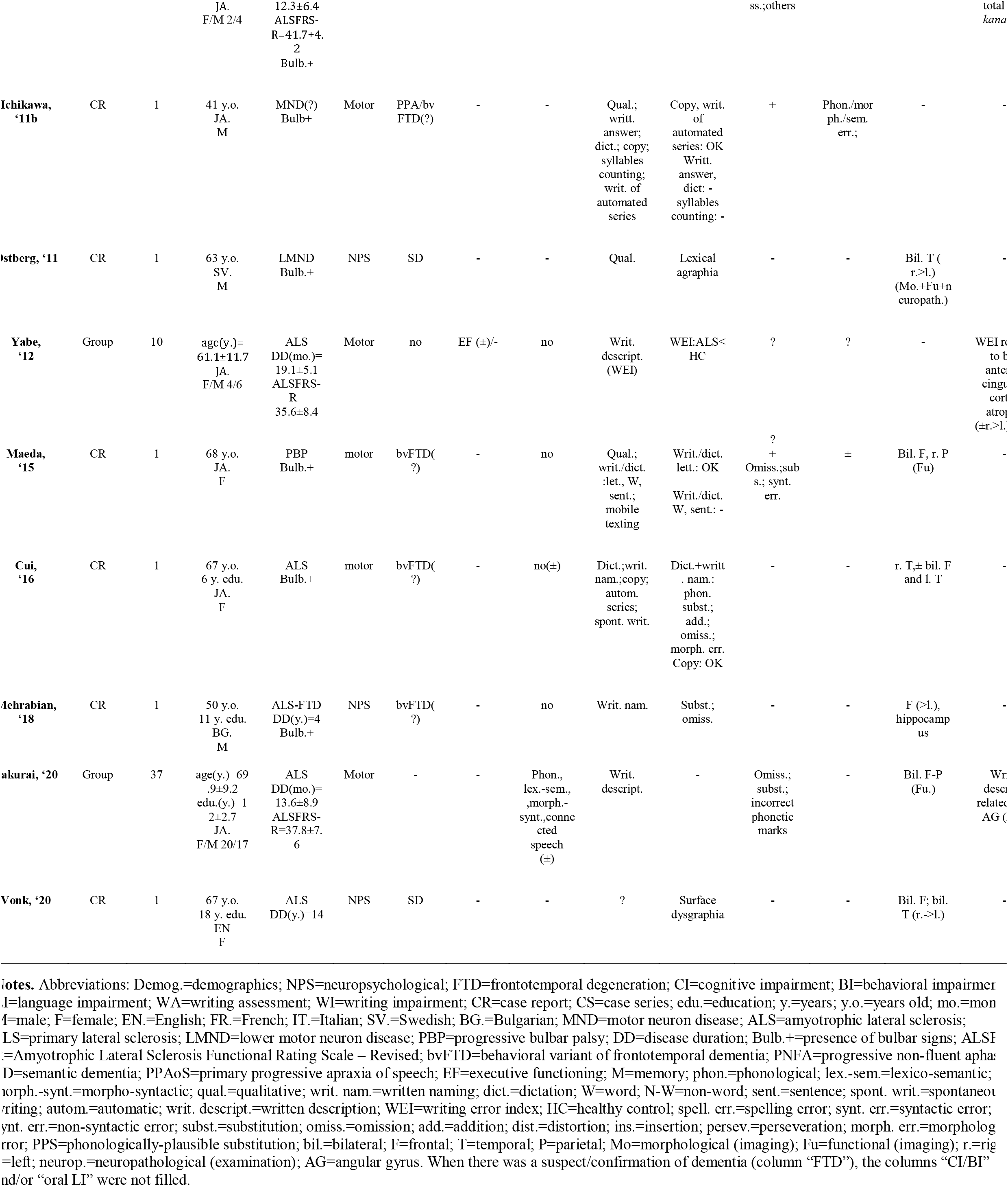
Summary of primary and secondary outcomes.

### 3.2. Group studies

#### 3.2.1. Group studies: western languages

Piquard *et al*. (2006) and Zago *et al*. (2008) assessed writing abilities in French and Italian primary lateral sclerosis (PLS) patients, respectively - comparing them to healthy controls (HC). Although both cohorts did not present with FTD or overt NPs impairment, central dysgraphic features were detected *via* writing-to-dictation tasks (of words/non-words/sentences) as either morpho-syntactic errors [Piquard *et al*., 2006] or phonetic/phonological paragraphias [Piquard *et al*., 2006; Zago *et al*., 2008]. Zago *et al*. (2008) also reported phonetic/phonological paragraphias in writing of non-words with preformed letters. The nature of writing deficits in Zago *et al*.’s (2008) contribution suggested a dysfunction of the graphemic buffer. Furthermore, the Authors found writing impairment to be associated with bulbar dysfunction – thus hypothesizing that spelling errors might have been accounted for by a rehearsal deficit due to the absence of motor feedback from the phono-articulatory system.

#### 3.2.2. Group studies: Japanese language

Tsuji-Akimoto *et al*. (2010) compared non-demented ALS patients to HCs on several NPs measures. Writing abilities were assessed *via* writing to dictation of *kana* and *kanji* characters, as well as by a writing errors index (WEI) computed from written description outputs. The WEI encompasses both phonetic/phonological and lexico-semantic paragraphias, as well as morpho-syntactic errors. At a group level, patients showed auditory comprehension and executive functioning (EF) deficits, as well as difficulties in writing *kanji* characters to dictation and a higher WEI than HCs. At an individual level, errors in writing to dictation of both *kana* and *kanji* characters were detected in patients. Interestingly, even those patients who did not show any errors when writing to dictation made both *kana* and *kanji* errors (of either phonetic/phonological, lexico-semantic or morpho-syntactic type) in written descriptions. Across tasks, the omission of *kana* letters (OKL) and a missing subject were found to be the most prevalent type of errors regarding phonetic/phonological and morpho-syntactic components, respectively. It is worth noting that the Authors described phonologically-plausible errors in both *kana* and *kanji* writing – thus suggesting surface dysgraphia. Moreover, the WEI was found to be mildly associated with disease duration. Yabe *et al*. (2012) again found that the WEI [Tsuji-Akimoto *et al*., 2010] was able to discriminate ALS patients from HCs, additionally showing that it was related to bilateral right-greater-than-left anterior cingulate cortex (ACC) dysfunction as assessed by positron emission tomography (PET). The Authors thus hint at EF deficits possibly accounting for writing errors at least to an extent. It is nonetheless worth noting that Yabe *et al*.’s (2012) cohort did present with mild only EF deficits. Recently, Sakurai *et al*. (2020) investigated the association between language functioning and neurofunctional measures (SPECT) in a cohort of 37 ALS patients. Phonological, lexico-semantic and morpho-syntactic dysfunctions, as well as reading and writing deficits were detected. Among different language measures, a written description task yielded the highest frequency of impairment (51.4%) and proved to be related to left both cortical and sub-cortical angular structures functioning - which are acknowledged to be involved in writing abilities [Sakurai *et al*., 2007]. When comparing ALS patients who displayed writing errors to those who did not, the former had lower educational attainment, reading scores as well as decremented inferior frontal, left supramarginal and bilateral parietal perfusion. Moreover, it is worth noting that age and age at onset were associated with writing errors.

### 3.3. Case series

#### 3.3.1. Case series: western languages

Ferguson & Boller (1977) described two non-demented English ALS patients showing phonetic/phonological paragraphias, morpho-syntactic errors and perseverations in spontaneous writing. Both patients developed bulbar signs. Although Case 1 was suspected to show also Gerstmann syndrome-like symptoms and presented with mild fronto-parietal neuropathological changes at *post-mortem* examination, Case 2 was not reported to present with overt NPs deficits. Case 1 also showed allographic preferences (uppercase) as the disease progressed. Duffy *et al*. (2006) similarly found spelling and morpho-syntactic errors in 4 out of 7 English MND patients – which nevertheless presented with apraxia of speech (AoS). Three out of the aforementioned 4 patients also presented with either bvFTD or PPA.

#### 3.3.2. Case series: Japanese language

Satoh *et al*. (2009) assessed writing abilities in 16 neuropsychologically-spared ALS patients by means of quantitative written language tests [Shewan & Kertsez, 1980; WAB Aphasia Test Construction Committee, 1986] and a task of counting of *morae* - *i*.*e*., the phonological units of *kana* words [Mishima *et al*., 2000]. *Kana* errors (*morae* miscounting and OKL) were found in 3 patients, whereas *kanji* errors in four; two patients simultaneously showed both *kana* and *kanji* errors. None of the patients showed morphological cortical abnormalities (magnetic resonance imaging, MRI), whereas single-photon emission computed tomography (SPECT) revealed bilateral frontal hypo-functioning in a patient presenting with both *kana* and *kanji* errors.

Ichikawa *et al*. (2008a) retrospectively assessed spontaneous writing samples and clinical features of 7 demented ALS patients. Patients presumably presented with bvFTD and showed anosognosia; 6 out of 7 and had bulbar-onset ALS. Writing errors were found in 6 patients: they showed both phonological/morphological *kanji* paragraphias (*i*.*e*., wrote a *kanji* character sharing either phonological or morphological similarities with the target) and *kana* errors (OKL, substitutions and perseverations); *kana-kanji* dissociations were also detected. Although oral LI could not be ruled out in 6 patients due to anarthria, it is worth noting that the patient whose oral output was intelligible and unimpaired still showed with *kanji* errors on a written description task. Bilateral frontotemporal atrophy (MRI) and hypo-functioning (SPECT) were detected in all patients – with a left predominance being found in three. The same group [Ichikawa *et al*., 2008b] similarly reported both *kana* and *kanji* errors in a retrospective cohort of bulbar-onset ALS patients, the majority of whom (10/15) either were demented (presumably presenting with bvFTD) or presented with BI. Thirteen out of 15 patients showed phonological/morphological *kana* paragraphias (including morpho-syntactic errors), whereas phonological/morphological *kanji* paragraphias were detected in 5 patients only; one patient showed only *kanji* errors. Furthermore, the Authors underlined that writing errors were also found in 2 non-anarthric patients who neither clinically showed oral LI nor CI/BI. The Authors subsequently described in more detail the 2 patients in a separate contribution [Ichikawa *et al*., 2010b]. Both patients presented with dysarthria. Phonological and lexico-semantic components of oral language were reported to be unimpaired, although both *kana* and *kanji* errors were detected both on a writing-to-dictation task and in spontaneous writing. Copy of letters was preserved; furthermore, a diagnosis of peripheral dysgraphia was ruled out. At structural and functional neuroimaging, left-greater-than right frontotemporal and parietal involvement was detected.

In a subsequent study, Ichikawa *et al*. (2010a) retrospectively investigated the association between writing errors and measures of cortical atrophy based on computer tomography (CT) [Evans, 1942; Frisoni *et al*., 2002] in 14 ALS patients. The majority of patients presented with bulbar/pseudobulbar signs at onset, and the presence of oral LI was either not detected or not assessable; seven patients showed bvFTD. Twelve patients showed *kana* errors, 8 of whom additionally presented with *kanji* errors – which nonetheless were not detected in isolation. The percentages of total and *kana* errors were found to be related to measures of frontal atrophy [Evans, 1942], whereas measures of temporal atrophy [Frisoni *et al*., 2002] were associated with *kanji* errors. Associations between the occurrence of bvFTD and writing errors were also reported. The same group further investigated the aforementioned anatomo-clinical associations also from a longitudinal standpoint [Ichikawa *et al*., 2011a]: writing abilities of six neuropsychologically-spared ALS patients were put in relation to two over-time-repeated CT-based measures of cortical atrophy. Five patients showed *kana* errors, 2 of whom also presented with *kanji* errors. Progression rate of rostral atrophy was found to be associated with both total and *kana* errors, whereas caudal atrophy proved not to be related to dsygraphic features; no associations were found between longitudinal atrophy measures and *kanji* errors separately. Anarthria prevented the Authors from stating whether oral LI was present at the time of the second CT measurement.

### 3.4. Case reports

#### 3.4.1. Case reports: western languages

Luchelli & Papagno (2005) described a patient presenting with primary progressive AoS who later developed MND. Overt NPs impairment/dementia was not detected. Phonetic/phonological paragraphias appeared to be highly frequent in several tasks; morpho-syntactic errors were also noted.

Östberg & Bogdanović (2011) described a Swedish patient presenting with semantic dementia (SD) who then developed lower motor neuron-predominant ALS. Severe impairment of semantic and lexical components was detected at oral language assessment: temporal right-greater-than-left abnormalities were detected at both morphological and functional neuroimaging, as well as at neuropathological examination. The Authors report that the patient showed both surface dyslexia and surface dysgraphia. Mehrabian *et al*. (2018) described a Bulgarian ALS-FTD patient carrying C9orf72 hexanucleotide repeat expansion who showed phonetic/phonological paragraphias on written naming. These dysgraphic features did reportedly worsen over time. The patient presented with multi-domain CI but was not classified as aphasic. MRI revealed bilateral left-greater-than-right frontal atrophy. Both the aforementioned cases showed bulbar signs. Vonk *et al*. (2020) recently reported a case of SD who reportedly developed ALS over a 14-years disease course. Among language dysfunctions, surface dysgraphia (along with surface dyslexia) was described. Although bilateral frontotemporal atrophy developed over time, in the first stages of the disease right-greater-than-left temporal atrophy was evident.

#### 3.4.2. Case reports: Japanese language

Dysgraphic features have been described in patients with MND and co-morbid PPA – either progressive non-fluent aphasia (PNFA) [Hyodo *et al*.,2002] or SD [Kito *et al*., 2010]. Hyodo *et al*.’s (2002) patient presented with PNFA and MND and reportedly showed phonetic/phonological paragraphias, as well as morpho-syntactic errors and perseverations in writing. Abnormalities were detected in left temporal and bilateral parietal cortices (MRI, SPECT). Kito *et al*. (2010) described a patient with SD who then developed MND. It is worth noting that difficulties in writing *kanji* were reported among the first cognitive symptoms. *Kana* errors appeared too, and dysgraphic features reportedly worsened over time. Bilateral left-greater-than-right frontal and rostro-temporal atrophy (MRI) and hypo-functioning (SPECT) was detected – and progressed over time. Both the aforementioned patients showed bulbar signs during the course of the disease.

Ishihara *et al*. (2010) described a neuropsychologically-impaired non-aphasic ALS patient who still reportedly showed mild-to-moderate phonological, lexico-semantic and morpho-syntactic impairment. Dysexecutive symptoms appeared to be prominent; bulbar signs were present. Right-greater-then-left rostro-temporal and bilateral frontal atrophy, as well as bilateral frontotemporal hypo-functioning, were detected at morphological (MRI) and functional (SPECT) neuroimaging, respectively. It is of great interest that particularly prominent histopathological abnormalities happened to be found in the caudal portions of the inferior frontal gyrus bilaterally – *i*.*e*., the so-called Exner’s area, which is acknowledged to be involved in writing abilities [Roux *et al*., 2009].

Ichikawa *et al*. (2011b) translated into English a 1893 Japanese case report [Watanabe, 1893], which described a patient presenting with bulbar palsy and progressive muscular atrophy who later developed dysgraphic features and disinhibited traits. Receptive language was reportedly preserved; severe dysarthria prevented the Author from assessing productive oral language. Both *kanji* and *kana* errors were reported – the latter happening to be more prevalent. Writing errors were detected both when the patient was asked to provide answers by writing and on writing-to-dictation tasks; copy and writing of automated series was preserved.

Maeda *et al*. (2015) reported a patient with progressive bulbar palsy who also presumably presented with bvFTD. Although severe EF deficits are described, the patient was reported to show neither overt oral LI nor dyslexia. The patient was able to write single *kana* characters, whereas presented with *kana* errors (OKL, substitution and morpho-syntactic errors) in writing words and sentences. The patient was reported to show *kana* phonetic/phonological paragraphias when writing text messages on her mobile phone. *Kanji* errors were nonetheless less frequent. Frontal atrophy/hypo-functioning was detected (MRI/SPECT); additionally, SPECT examination revealed right parietal hypo-functioning. Exner’s area was also reported to be involved (SPECT). The patient reportedly developed Gerstmann syndrome over the course of the disease.

Cui *et al*. (2016) described an ALS patient presumably presenting with bvFTD with evidence of frontal and right-greater-then-left temporal atrophy (MRI). Although multi-domain CI was present, oral language assessment did not reveal major deficits. With regard to written language, copy was spared, whereas phonetic/phonological and morphological errors were present in writing-to-dictation and written naming tasks.

## 4. Discussion

The present results suggest that central writing deficits may be a feature of MND patients’ cognitive dysfunction profile. Indeed, evidence suggestive of peripheral involvement appeared to be scarce, as well as, if present, to co-occur with central dysgraphic features [*e*.*g*., Ferguson & Boller, 1977]. Consistently, copying abilities often happened to be preserved [*e*.*g*., Ichikawa *et al*., 2011b]. However, it has to be noted that both central and peripheral involvement has been described in progressive dysgraphic syndromes [Graham *et al*., 1997] – this suggesting that a mixed peripheral/central underpinning should not be ruled out in MND patients either. Finally, it cannot be excluded that sthenic deficits in hand muscles might contribute to an extent to writing difficulties in this population [Cedarbaum *et al*., 1999].

Phonetic/phonological paragraphias appeared to be the most prevalent features of writing impairment in both eastern and western MND patients. Despite morpho-syntactic errors being also moderately frequent, they often happened to be due to spelling errors (*e*.*g*., *kana* postpositions in Japanese) [Tsuji-Akimoto *et al*., 2010]. Therefore, it can be hypothesized that the graphemic buffer might be involved – possibly along with the phonological route [Zago *et al*., 2008]. Although either a diagnosis of surface dysgraphia [*e*.*g*., Östberg & Bogdanović, 2011] or evidence suggestive of it (*e*.*g*., phonologically-plausible errors) [*e*.*g*., Tsuji-Akimoto *et al*., 2010], have been also reported, the lexico-semantic route appeared to be less involved than the phonological one. Moreover, although *kanji* errors (possibly due to lexico-semantic route impairment) have been also frequently reported in both demented and non-demented MND patients, *kana* errors (possibly due to phonological route/graphemic buffer impairment) have been more consistently detected.

*Extra*-linguistic predictors might also contribute to dysgraphic features in MND patients – in particular, those related to EF. Indeed, dysexecutive symptoms as well as frontal-type dementia have been proved to be associated/to co-occur with writing errors in MND patients [*e*.*g*., Ichikawa *et al*., 2010a; Tsuji-Akimoto *et al*. 2010]. Moreover, with regard to the nature of dysgraphic features, perseverations themselves might in turn hint at EF being impaired [*e*.*g*., Ferguson & Boller, 1977; Hyodo *et al*., 2002]. EF deficits as contributing to writing errors appears also to be supported as far as neural correlates are concerned – since frontal cortices often appeared to be involved in MND patients showing dysgraphic features [*e*.*g*., Yabe *et al*., 2012].

From a clinical standpoint, it is of interest to note that: 1) writing errors happened to be described in patients at relatively early stages of the disease (as far as disease duration and functional impairment are concerned) [*e*.*g*., Satoh *et al*., 2009; Tsuji-Akimoto *et al*., 2010; Sakurai *et al*., 2020]; 2) writing impairment has been found to progressively worsen over the course of the disease [*e*.*g*., Ichikawa *et al*., 2011]. Thereupon, writing errors might be regarded as an early marker of cognitive decline in MND patients.

The present review would provide empirical support for spelling tasks being included in MND-specific cognitive screening instruments [Woolley *et al*., 2010; Abrahams *et al*., 2014]. Moreover, since writing errors happened to be frequently detected by assessing writing samples, it might be useful to administer either spontaneous or non-spontaneous narrative writing tasks [*e*.*g*., Catricalà *et al*., 2017] in order to assess writing abilities in MND patients [Sakurai *et al*., 2020]. Writing-to-dictation tasks of words, non-words and sentences also appeared to be able to detect writing errors in MND patients - and thus might also be included [*e*.*g*., Zago *et al*., 2008].

Dysgraphic features have been frequently described in non-demented MND patients who either did not present with NPs impairment [*e*.*g*., Satoh *et al*.,2009] or showed mild CI/BI [*e*.*g*., Tsuji-Akimoto *et al*., 2010]. Similarly, writing impairment has been also reported in non-aphasic MND patients who either did not show oral LI [*e*.*g*., Piquard *et al*., 2006] or showed only mild language deficits [*e*.*g*., Zago *et al*., 2008]. With respect to this latter finding, it is worth mentioning that oral LI in non-aphasic patients was found not to mirror the nature of writing deficits [Tsuji-Akimoto *et al*., 2010]. Therefore writing deficits might represent a specific feature of MND patients’ profile [*e*.*g*., Ichikawa *et al*., 2008b]. This latter assertion also seems to be supported by evidence linking writing errors to abnormalities in cortical regions acknowledged to be specifically involved in writing (*e*.*g*., Exner’s area) [*e*.*g*., Ishihara *et al*., 2011].

Moreover, it is noteworthy that varying degrees of severity in writing impairment were reported – ranging from occasional writing errors in neuropsychologically-unimpaired patients [*e*.*g*., Satoh *et al*., 2009] to a full-blown dysgraphia in patients with co-morbid PPA [*e*.*g*., Hyodo *et al*., 2002]. This latter finding suggests that, in MND patients, dysgraphic features may occur along a *continuum* – similarly to other FTD-like NPs deficits [Strong *et al*., 2017]

Nonetheless, it has to be noted that left frontotemporal cortices have been systematically found to be involved in both demented and non-demented MND patients presenting with writing errors [see Tab. 1] – this hinting at perysilvian language networks abnormalities also possibly contributing to them [Purcell *et al*., 2011]. Interestingly, some contributions reported right-greater-than-left temporal involvement in neuropsychological-impaired/demented MND patients showing dysgraphia [Ishihara *et al*., 2011; Östberg & Bogdanović, 2011; Cui *et al*., 2016; Vonk *et al*., 2020]. As suggested by Cui *et al*.’s (2016), the latter finding might hint at dysgraphia possibly being a feature of MND patients presenting with the so-called right temporal variant of FTD [Coon *et al*., 2012; Ulugut Erkoyun *et al*., 2020].

Interestingly, parietal involvement along with suspected Gerstmann syndrome has been occasionally reported in MND patients presenting with peripheral/central dysgraphic features [Ferguson & Boller, 1977; Maeda *et al*., 2015; Sakurai *et al*., 2020]. It can be thus hypothesized that also cognitive processes relying on left parietal regions might contribute to writing deficits in this population [*e*.*g*., Sakurai *et al*., 2007].

The vast majority of contributions reported that MND patients showing dysgraphic features developed bulbar signs over the course of the disease. This may support the general notion that cognitive/language deficits in MND patients might be more frequent when neurodegeneration has spread to bulbar regions [Chiò *et al*., 2019; Shellikeri *et al*.,2019]. It can be postulated that to evaluate writing abilities may be particularly useful when assessing bulbar-plus MND patients’ cognitive/language profile. Moreover, writing also represents the only modality through which both language and several cognitive functions can be assessed in severely dysarthric/anarthric patients. As previously mentioned [see 2.3], this latter assertion can as well be regarded as a limitation of the present review – since it might have led to an overestimation of the prevalence of writing disorders in this population. On the other hand, for the same reason, severe dysarthria/anarthria might have led to an underestimation of co-morbid NPs deficits [see 2.3].

It may be worth noting that the vast majority of the studies that were included reported dysgraphic features in MND phenotypes involving either only upper or both upper and lower motor neurons (*e*.*g*., PLS and ALS). Only one study reported dysgraphia in a patient affected with SD and co-morbid LMND [Östberg & Bogdanović, 2011]. It can be therefore hypothesized that writing deficits, as well as cognitive impairment in general, might be more prevalent when upper motor neuron dysfunction is present - due to a greater degree of cortical involvement [Aiello *et al*., 2020].

The present work is of course not free of limitations. First, as previously stated [see 2.3], it is challenging to compare western to Japanese taxonomy when it comes to dysgraphia [Purcell *et al*., 2011; Sakurai, 2019]. Therefore, the potential lack of homogeneity on categorizing errors might have led to distortion in interpreting results, as well as in drawing cross-linguistically valid inferences. Furthermore, it has to be noted that the majority of the studies that were included were case reports/case series – this implying that a comparison with HCs often lacked, as well as that high levels of between-studies variability were present with respect to both adopted methods and reported elements.

## 5. Conclusions

Central dysgraphic features were reported in both neuropsychologically-impaired and – unimpaired MND patients. By contrast, evidence of peripheral dysgraphia was scarce. Phonetic/phonological paragraphias and morpho-syntactic errors were frequently reported. Although FTD was frequently co-occurring, neither cognitive nor language impairment fully accounted for writing impairment in some patients. Patients displaying writing deficits often presented with bulbar signs and left perisylvian cortices involvement (including Exner’s area and the left angular gyrus). Writing deficits proved to be associated with abnormalities in executive functioning and their neural substrates. Writing-to-dictation tasks as well as writing samples assessment proved to be useful to detect writing errors.

Dysgraphic features in MND patients are likely to be due to dysfunctions of the graphemic buffer/the phonological route. The lexico-semantic route appeared to be less involved. However, a mixed peripheral/central involvement cannot be ruled out. In this population, executive/attentive deficits are likely to contribute to writing errors as well. Writing deficits might thus be specific of MND patients’ cognitive/language impairment profile. The evaluation writing abilities through writing-to-dictation/narrative writing tasks may be useful when assessing cognition/language in both neuropsychological-impaired and -unimpaired MND patients - especially when severe dysarthria/anarthria is present.

## Data Availability

No datasets are associated to this manuscript.

